# Associations between measures of social distancing and SARS-CoV-2 seropositivity: a nationwide population-based study in the Netherlands

**DOI:** 10.1101/2021.02.10.21251477

**Authors:** Eric R.A. Vos, Michiel van Boven, Gerco den Hartog, Jantien A. Backer, Don Klinkenberg, Cheyenne C.E. van Hagen, Hendriek Boshuizen, Rob S. van Binnendijk, Liesbeth Mollema, Fiona R.M. van der Klis, Hester E. de Melker

**Affiliations:** Centre for Infectious Disease Control, National Institute for Public Health and the Environment (RIVM), Antonie van Leeuwenhoeklaan 9, 3720 MA Bilthoven, the Netherlands

**Keywords:** COVID-19 pandemic, SARS-CoV-2 seroprevalence, social distancing, transmission, the Netherlands

## Abstract

This large nationwide population-based seroepidemiological study provides evidence on the effectiveness of physical distancing (>1.5m) and indoor group size reductions on SARS-CoV-2 infection. Additionally, young adults seem to play a significant role in viral spread, opposed to children up until the primary school age with whom close contact is permitted.

## INTRODUCTION

The coronavirus disease 2019 (COVID-19) pandemic is an unprecedented global crisis. Stringent measures to suppress the spread of its causative agent severe acute respiratory syndrome coronavirus-2 (SARS-CoV-2) have been implemented to reduce incidence of disease and prevent health systems from becoming overwhelmed. Assessment of the impact of social-distancing measures is vital for informing public health decisions, particularly since the worldwide availability of vaccines is still very limited in this phase.

In the Netherlands, the first case of COVID-19 was reported on February 27, 2020. Key governmental interventions implemented since mid-March, 2020, included: keeping physical distance (≥1.5m) for those aged >12 years, whereas close contact between 13-17 year-olds was permitted; closure of schools, restaurants/bars/cafés, cultural institutions, sport facilities; working from home if possible; prohibition of contact professions; closure of nursing homes to visitors; and reducing group sizes. In May, primary schools and daycare were re-opened, and contact professions were allowed to resume. From June onwards, measures were further relaxed, while adhering to physical distancing measures.

Seroprevalence of antibodies against SARS-CoV-2, acquired from validated laboratory assays and well-designed population-based studies, provide an unbiased indicator of cumulative infection [1, 2]. In combining seroprevalence with questionnaire data, the current nationwide population-based study (PIENTER-Corona, PICO) [3] – set-up after the first epidemic wave in the Netherlands in June, 2020 – enabled us to identify risk factors for infection to support assessment of the impact of globally-applied social distancing measures.

## METHODS

Randomly-selected participants of all age groups from the first PICO-serosurvey in April, 2020 [3, 4], were invited for the current study in June and 2,317 enrolled. To enhance countrywide geographical coverage, and given the low anticipated seroprevalence, this cohort was supplemented with an additional sample of 4,496 randomly sampled participants (n_total_=6,813) (**Supplement–p3-4**). Participants were requested to collect a fingerstick blood sample and return it by mail. An (online) questionnaire was completed on potential SARS-CoV-2 exposure (number and age group of non-household close contacts (<1.5m) the day before filling out the questionnaire, attendance of indoor meetings with >20 persons, nursing home visits, working from home last week, profession, close contact (voluntary) work with patients/clients and children, and household size); and sociodemographic characteristics (sex, age, ethnic background, religion, educational level, postal codes were used to determine geographical sites).

Quantitative measures of serum IgG antibodies against SARS-CoV-2 Spike-S1 antigen were derived via a validated multiplex-immunoassay [5]. Based on low anticipated seroprevalence [3], we aimed for a specificity of 99.9% to keep false positive rates to a minimum. Mixture model analyses (using a validation panel as prior distribution) showed that such specificity could be obtained (at a cut-off for seropositivity of 0.04 log(AU)/mL) with associated sensitivity of 94.3% (95% confidence interval (CI) 90.6-96.7) (**Supplement–p6-15**). Applying this cut-off, all seroprevalence estimates (and 95% CIs) for the general Dutch population took into account the survey design, included weighting factors to match the distribution of the general Dutch population (based on sex, age, ethnic background and degree of urbanization; **Supplement–p6**), and were controlled for test characteristics subsequently [6, 7]. Smooth age-specific seroprevalence was modelled with B-splines (second degree, three percentile-placed internal knots, following lowest Akaike’s Information Criteria (AIC)).

Risk factors for seropositivity were identified using random-effects logistic regression – taking into account municipality as a unit of clustering. In the main analysis, all participants without missing data for the tested determinants were included (n=6,331). Odds ratios (OR) and 95% CIs were derived from univariable analyses, and two-way interactions with age were tested for significance. Variables with an overall p<0.15 were tested in multivariable analysis, in which stepwise-backward selection was applied yielding a final model including only independent risk factors (based on lowest AIC). Sensitivity analyses were performed applying forward selection; and by testing models without religion (n=6,487) – as this variable comprised the most missing values – without educational level (n=6,339) and without non-household contact data (n=6,338) – the latter two to test potential associations with profession.

Analyses were performed using Stan v.2.21 (mixture modelling), and SAS v.9.4 (SAS Institute Inc., USA). The study was approved by the Medical Ethics Committee MEC-U (Clinical Trial Registration NTR8473) and all participants provided written informed consent.

## RESULTS

Median inclusion date was June 14, 2020 (range: June 9–Augustus 24; 90% was enrolled by June 22) (note: sociodemographic characteristics available from non-responders were compared to responders and shown in **Supplement–p4-5**). The cohort comprised 55% women and regions were equally represented following population size (**Supplement–p5-7**). Half of the participants reported to have had ≥2 non-household close contacts the day before filling out the questionnaire. Since the start of the epidemic, one quarter had attended an indoor meeting with >20 persons, and 8% had visited a nursing home. Among 18-66 year-olds, 36% (voluntarily) worked in close contact with clients/patients, 18% was a healthcare worker, and 40% had been (partly) working from home last week.

After the first epidemic wave, overall seroprevalence in the Dutch population was 4.5% (95% CI 3.8-5.2). No difference was observed between sexes and ethnic backgrounds. Estimates were low (0-2%) in children aged 1-12 years, high (9%) in young adults in their early twenties, and 4-7% in individuals ≥35 years (**Figure1A**). Low urbanized areas were hit hardest, predominantly in the South-East (up till 16%) (**Supplement–p16**).

**Figure 1:**
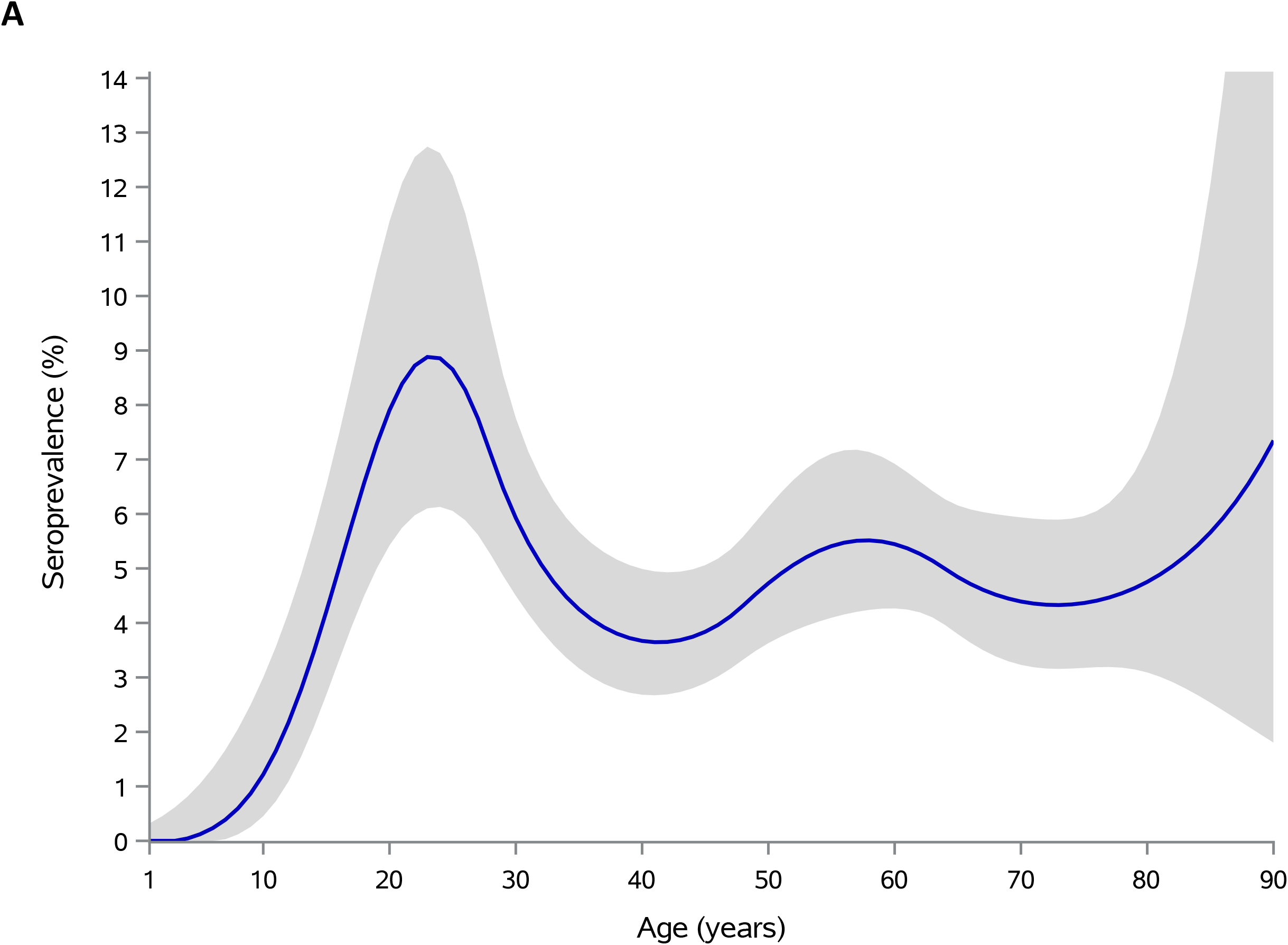
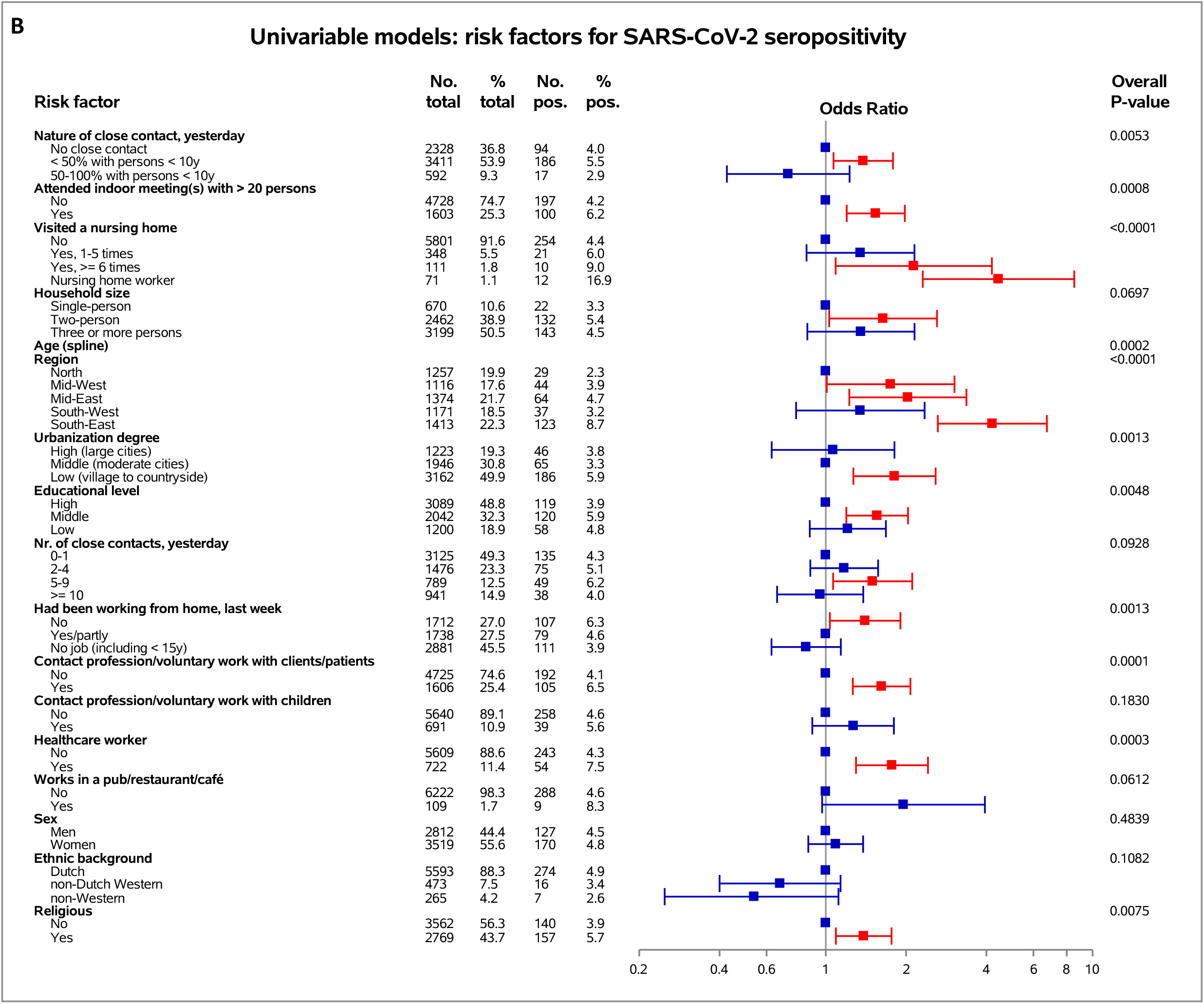
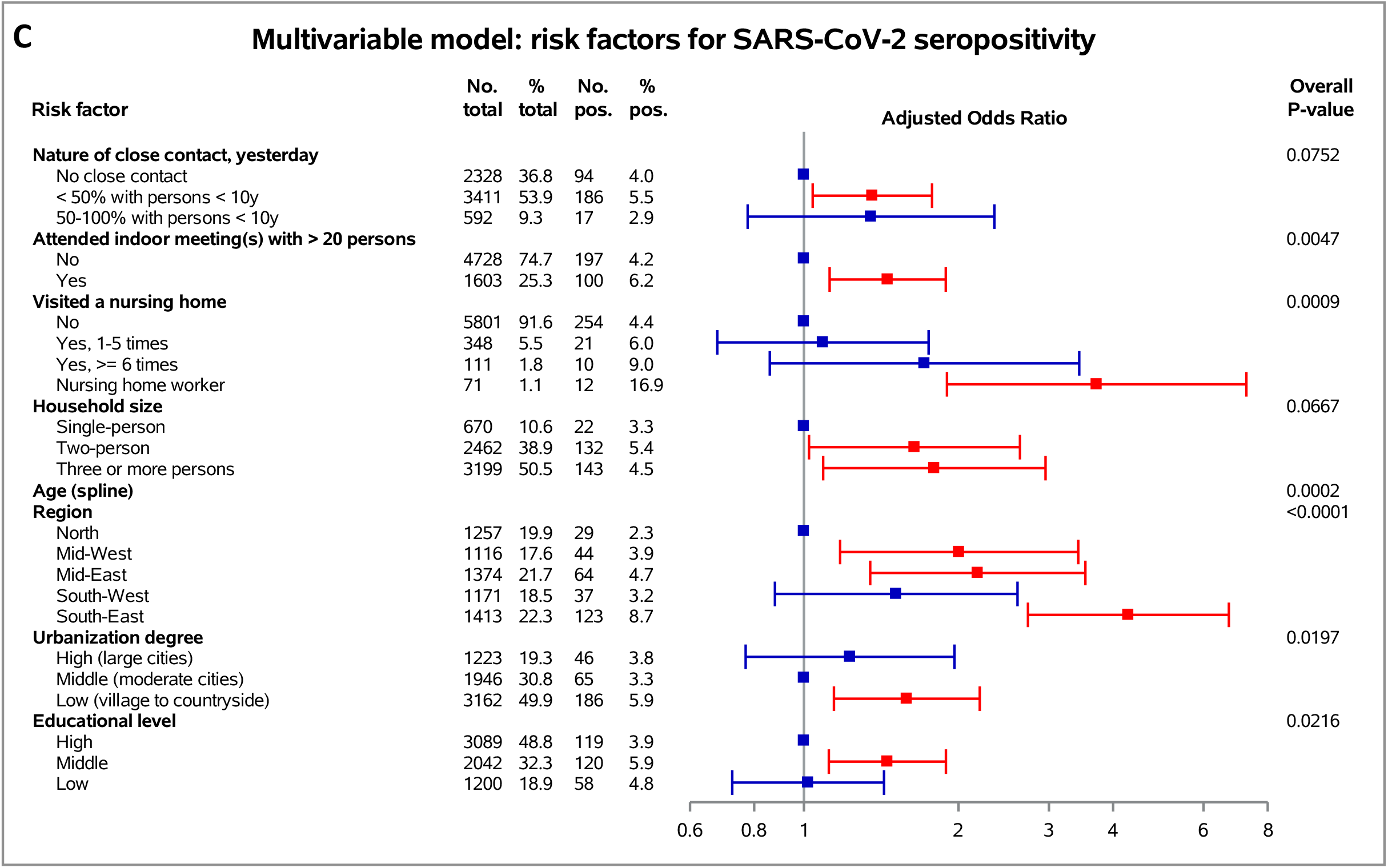
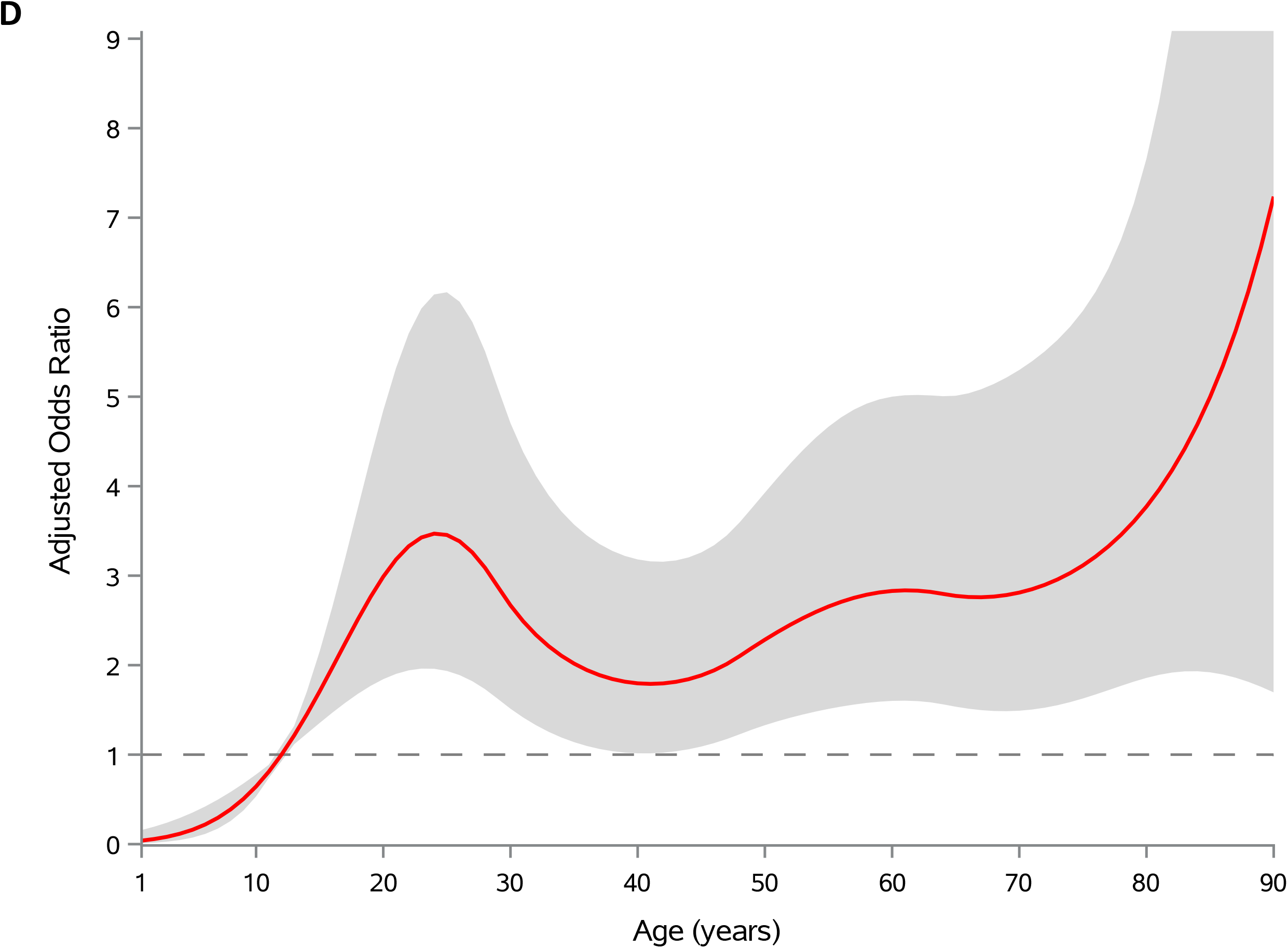
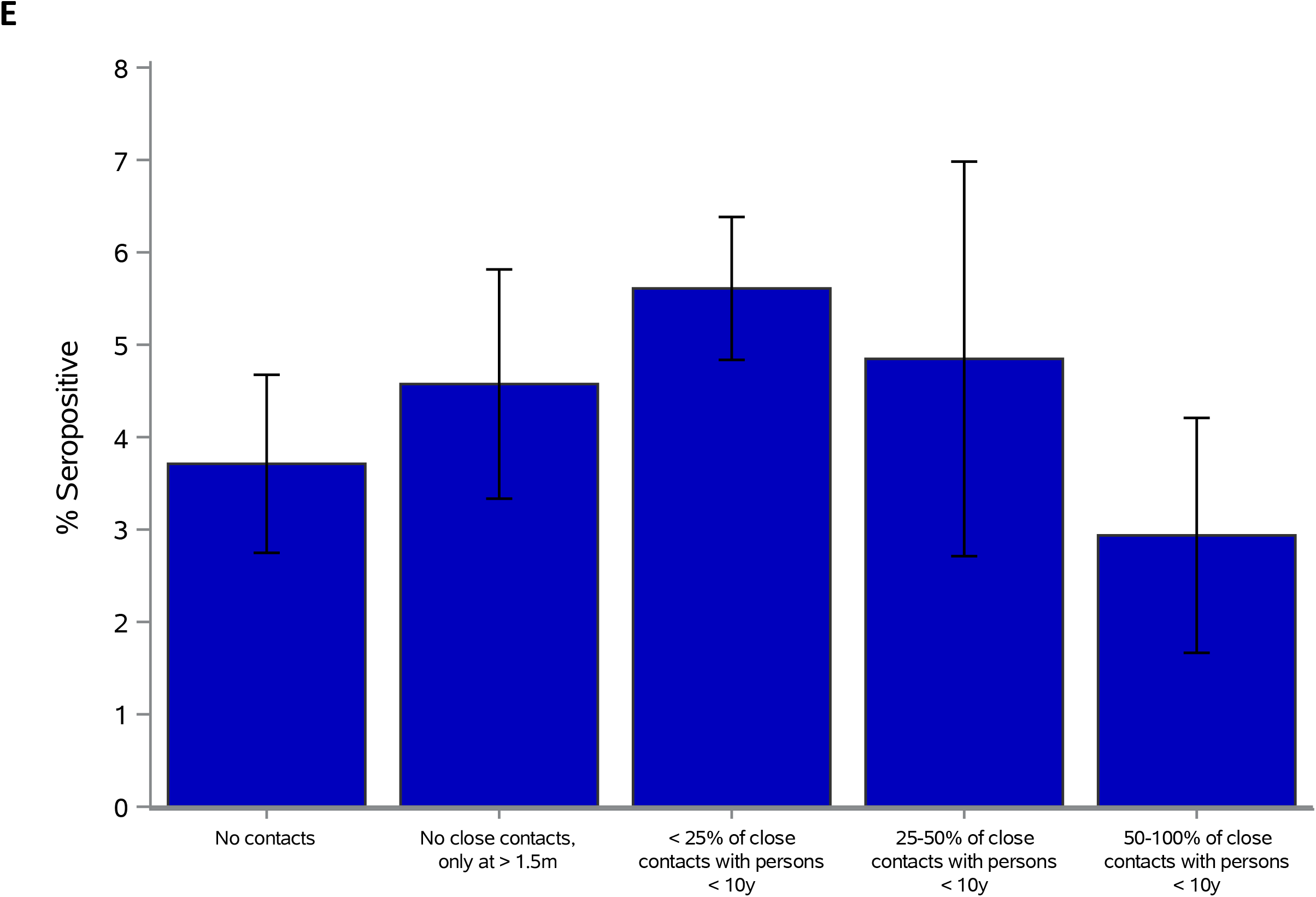
**A**. Shows the weighted smooth age-specific SARS-CoV-2 seroprevalence with 95% confidence envelope in the general population of the Netherlands after the first epidemic wave. **B and C** display the risk factor analyses for SARS-CoV-2 seropositivity. Number (and %) of total participants per potential risk factor-category are provided as well as the number (and %) of seropositive participants (pos.), and overall P-values. Forest plots are shown of crude odds ratios (OR) for univariable analyses (**B**) and adjusted odds ratios (aOR) for the multivariable analysis (**C**), and depicted by squares and 95% confidence intervals (95% CIs) with lines: those in red are significantly associated with seropositivity and those in blue are non-significant. Age was included with a flexible (spline) function. Time window of attending indoor meetings with > 20 persons and visiting a nursing home concerned the beginning of the epidemic in the Netherlands (February 27, 2020) until the day of filling out the questionnaire or until closure (for visitors) of nursing homes (March 20, 2020), respectively. Nature and number of non-household close contacts yesterday, and working from home last week, concerned the day or week, respectively, before filling out the questionnaire. Receiver operating characteristic curve analysis of the multivariable model yielded an area under the curve (as a measure of goodness-of-fit) of 0.72. **D**. shows the aOR with 95% confidence envelope for age derived from the multivariable model, with 12 year as reference category. **E**. displays SARS-CoV-2 seropositivity (and 95% confidence intervals) by number and nature of non-household close contact the day before filling out the questionnaire. Nature of non-household close contact was defined as the proportion of non-household close contacts with children aged < 10 years of the total number of non-household close contacts.

All potential risk factors for seropositivity tested in univariable analyses are shown in **Figure1B** (and for age in **Supplement–p16**). Close contact (voluntary) work with children was not associated. Also, work with clients/patients and total number of non-household close contacts did not remain in the final model. Social distancing-related risk factors in the multivariable model included (**Figure1C**): non-household close contacts with ≥50% persons ≥10 years as compared to no contacts, whereas close contact with ≥50% children aged <10 years was not statistically significantly associated (see also **Figure1E**); attending indoor meetings with >20 persons; working in a nursing home (rather than a visitor), increased household size; and age, with low adjusted odds in children ≤12 years, whilst over 2.5 times higher odds in adults aged 18-30 and ≥50 years as compared to 12-year-olds (**Figure1D**). Sensitivity analyses yielded similar results.

## DISCUSSION

Here, we provide evidence from a large population-based study on the effectiveness of physical distancing (>1.5m) as well as indoor group size reductions on SARS-CoV-2 infection, and these data substantiate policy of allowing close contact between teachers and children in primary school.

Our results on physical distancing are in line with the few previous reports mostly derived from healthcare settings and households [8]. Seroprevalence rates were low in children aged ≤12 years despite close contact, and similar to observations from other European countries with comparable nationwide estimates [1, 9]. Interestingly, the likelihood of infection among persons in close contact with children was not statistically significantly increased, most likely indicating a low contribution in transmission, as suggested previously [10-13]. On the other hand, particularly young adults, who engage in relatively more social interaction as opposed to older age groups [14] and often living in larger (student) households, most probably play an increased role. Applying physical distancing measures within households may not always be feasible, however stressing its relevance in outbreak management could help to reduce (ongoing) transmission. Further, like in ample other countries [15], these data underline the increased risk of infection among nursing home workers. Hence, while working with the most vulnerable, this requires specific attention.

Our study has strengths and limitations. Strength is that our study provides a large population sample covering a full age-range from young to old, combining a sound indicator of prior infection, i.e., seropositivity, to extensive questionnaire data. Also, samples could be classified accurately since antibodies were measured with a highly specific and sensitive immunoassay. Limitations include the relatively low response rate, which might have introduced potential selection bias, e.g., of relatively more health-conscious individuals adhering to social distancing measures. Further, some variables might be proxies of risk of viral exposure, e.g., on contacts, thus associations should be interpreted with care as they may not reflect causal effects.

In conclusion, these results underscore the effectiveness of the social distancing-related measures to reduce SARS-CoV-2 transmission in an era of limited availability of vaccines. Additionally, our data suggest a diminished role of young children in viral spread, which may justify decisions to keep primary schools open, while young adults seem to play a more substantial role.

## Data Availability

Our data are accessible to researchers after publication and upon reasonable request for data sharing to the principal investigator

## ACKNOWLEDGMENTS

We gratefully acknowledge the participants of the current study. Moreover, this study would not have been possible without the instrumental contribution of colleagues from the National Institute of Public Health and Environment (RIVM), more specially the department of Immunology of Infectious Diseases and Vaccines (IIV) (regarding logistics and laboratory analyses), and the department of Epidemiology and Surveillance (EPI) (concerning logistics, and methodological and statistical insights). Furthermore, we would also like to thank Maarten Schipper for performing the sampling, and Susan van den Hof (EPI department head) for reviewing the manuscript.

## ETHICAL APPROVAL

This study protocol was approved by the Medical Ethics Committee MEC-U, the Netherlands (Clinical Trial Registration NTR8473), and conformed to the principles embodied in the Declaration of Helsinki.

## Supplementary Information

### 1 Introduction

In this supplement we detail our sampling strategy, provide information on non-response rates, and explain how we have included post-stratification weighting in the analyses. The risk factor analyses in the main text use logistic regression based on binary classification of the data (seronegative versus seropositive). Here, we also provide an underpinning of this classification using a two-component mixture model. In this model, samples are not rigidly classified as either seronegative or seropositive, but belong to either the negative or positive component with certain probability [1, 2]. As the probability of seropositivity may depend on age, we model the mixing parameter (i.e. the probability of seropositivity, or seroprevalence) with an age-dependent penalized spline [3]. We fit the model to antibody concentration measurements from the population sample described in the main text while incorporating information from a test panel of proven negative and positive samples [4]. Subsequently, we derive test characteristics (sensitivity, specificity) for various cut-offs, showing that the binary classification used in the main text performs well. In a final step we present additional results for the weighted seropositivity estimates by municipal health services, and for the age-specific odds of seropositivity.

### 2 Sampling

The current cohort includes persons who had participated in our earlier SARS-CoV-2 serosurveillance study in March, 2020 (sample 1) and an additional nationwide sample (sample 2). Details on the first sample have been described previously [5, 6]. In this previous study, 2, 634 participants had been included. Anticipating a 10% drop-out rate from the first study, and given the low estimated seroprevalence in the first study (2.8%), we aimed to increase the overall power of the study. Hence, the initial cohort was supplemented with an additional sample of randomly sampled participants from the Dutch population registry as of May, 2020.

For this second sample, persons were randomly drawn from five regions with roughly similar population size (North: provinces of Groningen, Friesland, Drenthe and Overijssel; Mid-West: provinces of Flevoland and Noord-Holland; Mid-East: provinces of Gelderland and Utrecht; South-West: provinces of Zuid-Holland and Zeeland; South-East: provinces of Noord-Brabant and Limburg), and from 17 pre-defined age groups (1-4, 5-9, 10-14, 15-19, 20-24, 25-29, 30-34, 35-39, 40-44, 45-49, 50-54, 55-59, 60-64, 65-69, 70-74, 75-79, 80-89 years).

A total sample size of 6, 400 participants, i.e. with an average of 380 participants per age group, would enable us to estimate an overall and age-specific seroprevalence with a precision of 1.25% and 5%, respectively. Following previous experience, we anticipate a response rate of at least 15%. Hence, for the additional sample, we randomly selected 27, 200 persons from the population registry, of which 26, 854 remained eligible for participation after an initial screening.

### 3 Non-response and weighting

All randomly-selected persons who were invited in the first serological study were also invited for the current study. Of these, 2, 317 participated in the current study. This cohort was subsequently supplemented with an additional sample (as described above). Specifically, we invited 26, 854 randomly-selected persons of which 4, 496 participated, resulting in an overall number of 6, 813 participants. Table S1 shows the number of participants and response rates, stratified by sex, age group, region, and ethnic background. See main text for details and discussion.

**Table S1.** Overview of the study data. Ethnicity is missing for 16 invited persons.

|  |  | Non-responder |  | Responder |  | Total |
| --- | --- | --- | --- | --- | --- | --- |
|  |  | N | % | N | % | 31,780 |
| Total |  | 24,967 | 78.6 | 6,813 | 21.4 |  |
| Sex | Man | 12,609 | 50.5 | 3,042 | 44.7 | 15,561 |
|  | Women | 12,358 | 49.5 | 3,771 | 55.4 | 16,129 |
| Age | 1-4 | 1,740 | 7.0 | 220 | 3.2 | 1,960 |
|  | 5-9 | 1,637 | 6.6 | 285 | 4.2 | 1,922 |
|  | 10-14 | 1,567 | 6.3 | 319 | 4.7 | 1,886 |
|  | 15-19 | 1,591 | 3.4 | 304 | 4.5 | 1,895 |
|  | 20-24 | 1,542 | 6.2 | 300 | 4.4 | 1,842 |
|  | 25-29 | 1,779 | 7.1 | 398 | 5.8 | 2,177 |
|  | 30-34 | 1,293 | 5.2 | 369 | 5.4 | 1,662 |
|  | 35-39 | 1,519 | 6.1 | 408 | 6.0 | 1,927 |
|  | 40-44 | 1,439 | 5.8 | 448 | 6.6 | 1,887 |
|  | 45-49 | 1,423 | 5.7 | 457 | 6.7 | 1,880 |
|  | 50-54 | 1,365 | 5.5 | 548 | 8.0 | 1,913 |
|  | 55-59 | 1,323 | 5.3 | 544 | 8.0 | 1,867 |
|  | 60-64 | 1,226 | 4.9 | 591 | 8.7 | 1,817 |
|  | 65-69 | 1,250 | 5.0 | 626 | 9.2 | 1,876 |
|  | 70-74 | 1,326 | 5.3 | 501 | 7.4 | 1,827 |
|  | 75-79 | 1,410 | 5.7 | 134 | 2.0 | 1,752 |
|  | 80-90 | 1,537 | 6.2 | 153 | 2.3 | 1,690 |
| Region | North | 5,029 | 20.1 | 1,357 | 19.9 | 6,386 |
|  | Mid-West | 4,957 | 19.9 | 1,211 | 17.8 | 6,168 |
|  | Mid-East | 4,825 | 19.3 | 1,469 | 21.6 | 6,294 |
|  | South-West | 5,060 | 20.3 | 1,248 | 18.3 | 6,308 |
|  | South-East | 5,096 | 20.4 | 1,528 | 22.4 | 6,624 |
| Urbanization degree | High | 6,038 | 24.2 | 1,319 | 19.4 | 7,357 |
|  | Middle | 7,670 | 30.7 | 2,101 | 30.8 | 9,771 |
|  | Low | 11,259 | 45.1 | 3,393 | 49.8 | 14,652 |
| Ethnic background | Dutch | 18,598 | 74.5 | 5,996 | 88.0 | 24,594 |
|  | Non-Dutch Western | 2,389 | 9.6 | 512 | 7.5 | 2,901 |
|  | Non-Western | 3,964 | 15.9 | 305 | 4.5 | 4,269 |

Post-stratification weights are assigned to each participant to standardize seroprevalence estimates, using census data of the Netherlands from January 2020. Since our cohort consists of two samples, weights are calculated for each sample separately. Per study sample, weights are assigned to each participant based on their membership to specific census strata (in total 112): for Dutch ethnic background, strata are designed for age group (1-4, 5-9, 10-14, 15-19, 20-24, 25-29, 30-34, 35-39, 40-44, 45-49, 50-54, 55-59, 60-64, 65-69, 70-74, 75-90 years), urbanization level (high, middle, low), and sex; and for other ethnicity groups strata were based on age group (1-9, 10-34, 35-59, 60-90 years) and sex.

Subsequently, post-stratification weights are defined as the proportion of each stratum represented in the Dutch population divided by the analogous proportion in the study sample. Specifically, participant weights *w*_*ij*_ for participants in stratum *i* and study *j* are calculated as

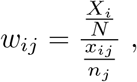

where *X*_*i*_ is the total number of persons in stratum *i, N* is the total population size (i.e. the Netherlands), *x*_*ij*_ is the number of participants in stratum *i* in study sample *j*, and *n*_*j*_ is number of participants in sample *j*.

### 4 Data

Figure S1 shows the regional distribution of samples, and Figure S2 shows the antibody concentration measurements by age. For the analyses, we also include a validation panel that has been used for validation of the assay [4]. Specifically, we take a set of 384 samples from uninfected persons that had been drawn from the Dutch population before the pandemic, and a set of 115 proven SARS-CoV-2 infections with mild to severe disease [4]. Mean and standard deviation of the (log-transformed) measurements are *µ*_uninfected_ = −2.3 (arbitrary units) and *σ*_uninfected_ = 1.0 for the uninfected group, and *µ*_infected_ = 3.0 and *σ*_infected_ = 2.1 for the infected group.

**Figure S1.**
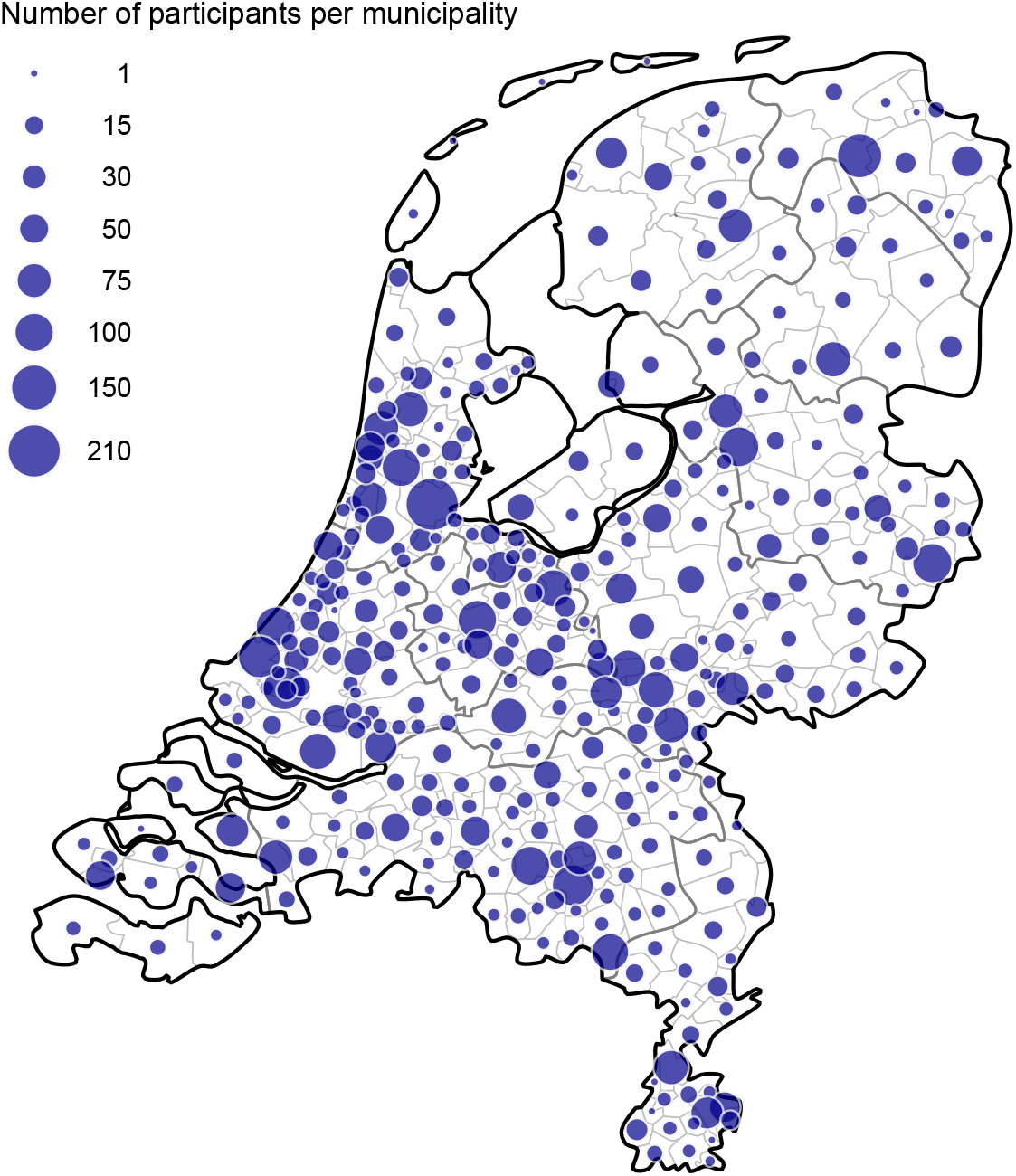
Regional distribution of samples. Notice that the western part of the Netherlands is the most densely populated area and also has large number of samples, thus attaining good population coverage.

### 5 Mixture model

Survey participants are assumed to be either seropositive or seronegative. These two classes are characterized by distributions for antibody measurements, denoted by *f*_neg_ and *f*_pos_ and specified by parameters *θ*_neg_ and *θ*_pos_. Further, the mixing parameter (probability of seropositivity) depends on age and is denoted by *p* (*a*). For *n* = 6, 813 participants, the set of participant ages and observed measurements are given by **a** = (*a*_*k*_) and **x** = (*x*_*k*_) (*k* = 1, …, *n*), respectively. Throughout we use normal distributions for the components of the mixture of the log-transformed data, so that *θ*_neg_ = (*µ*_neg_, *σ*_neg_) and *θ*_pos_ = (*µ*_pos_, *σ*_pos_), while the mixing parameter is modelled with a Bayesian penalized-spline using cubic basis functions and first order penalization [7, 8]. Throughout, we consider the age range [0, 100] years, placing knots at 10-year intervals (11 knots in total), so that the total number of basis functions is 13 [7, 8].

**Figure S2.**
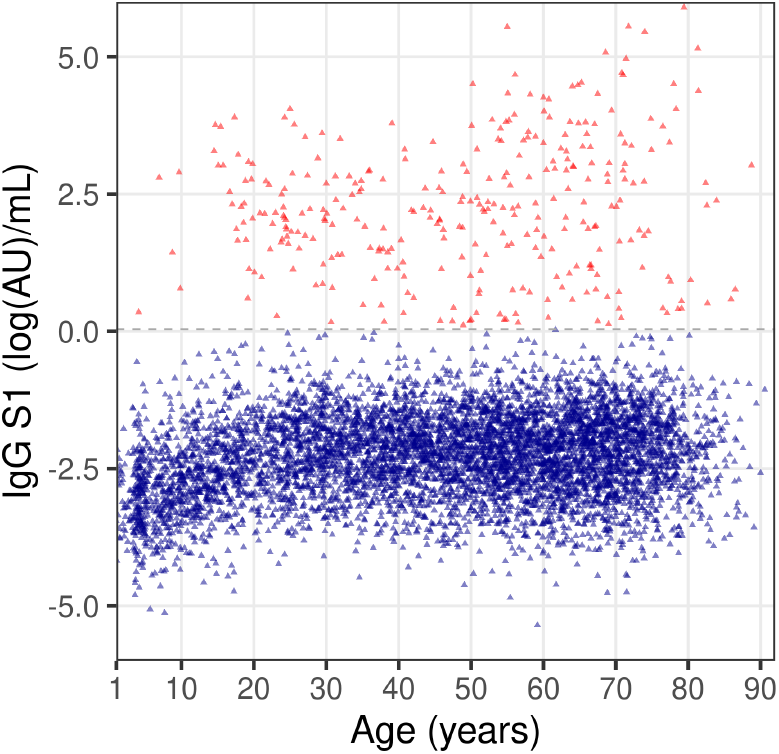
Overview of the data. Shown are (log-transformed) antibody concentrations of all 6,813 samples in the national sample as function of age. Here, samples are classified as seronegative below the cut-off of 0.04 (log(AU)/mL)(blue) and as seropositive above the cut-off (red).

### 6 Estimation

Parameters are estimated in a Bayesian framework using Hamiltonian Monte Carlo, implemented in Stan [9]. To improve performance at low prevalence, we employ a logistic transformation for the age-specific prevalence.

Prior distributions for the means and standard deviations of the seronegative and seropositive components are based on the uninfected and infected samples from the validation set. As the uninfected set is obtained from random samples from the Dutch population in 2006/2007 and 2016/2017 as well as a panel comprising cases with influenza-like illness, and the seropositive set contains cases with symptoms of disease and may be less representative of cases in the population, we take informative prior distributions for the parameters of the seronegative component, a weakly informative prior distribution for the mean of the seropositive component, and provide no explicit prior distribution for the standard deviation of the seropositive component. Specifically, we take

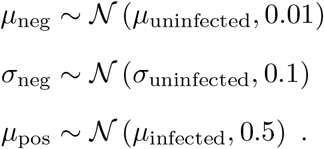

For the spline smoothing parameter (RWvar) we take an inverse gamma distribution [8],

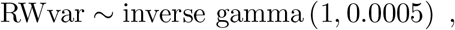

and for the weights of the spline base functions *w*_*i*_ (*i* = 1 … 13), we take

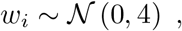

where it should be noted that the prior weights are defined on the logistic scale.

**Table S2.** Parameter estimates (selected posterior quantiles) with selected convergence diagnostics.

| Parameter | $\widehat{R}$ | $n_{\text{eff}}$ | 2.5% | 50% | 97.5% |
| --- | --- | --- | --- | --- | --- |
| $\mu_{\text{neg}}$ | 0.997 | 1071 | -2.311 | -2.297 | -2.284 |
| $\sigma_{\text{neg}}$ | 0.997 | 964 | 0.742 | 0.756 | 0.770 |
| $\mu_{\text{pos}}$ | 0.996 | 1066 | 1.967 | 2.168 | 2.336 |
| $\sigma_{\text{pos}}$ | 1.003 | 1126 | 1.216 | 1.339 | 1.501 |
| RWvar | 1.000 | 1030 | 0.008 | 0.042 | 0.169 |

Results from properly converged chains are obtained within hours (using 10 cores on our servers). Estimates for the parameters defining the mixing distribution and the spline smoothing parameter are given in Table S2, together with convergence diagnostics 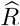 and *n*_eff_ [9]. In a sensitivity analysis we have re-run the fitting procedure with uninformative prior distributions (only assuming that *µ*_pos_ *> µ*_neg_). These analyses yield virtually identical results (not shown).

**Figure S3.**
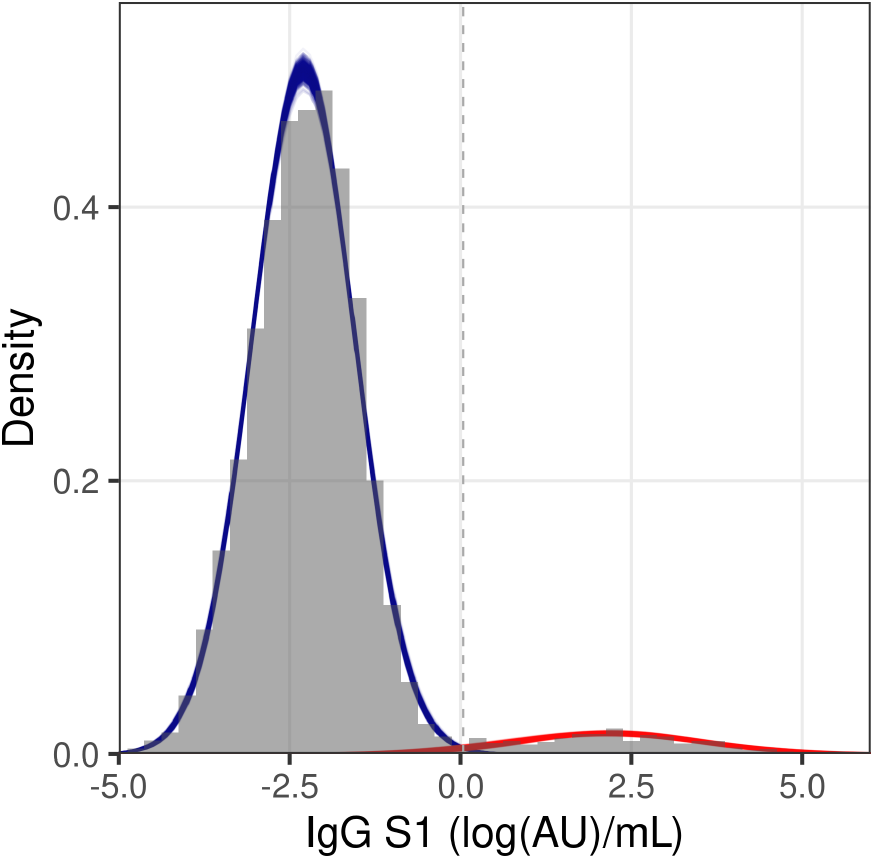
Data and model fit. Shown are the data (gray histograms) and fit of the mixture model (blue: seronegative component; red: seropositive component). The age-specific prevalence is modelled with a penalized spline, and the mixing distributions are weighted with the overall posterior probability of infection. Shown are 1, 000 samples from the posterior distribution.

Figure S3 gives a visualisation of the data (gray histograms) and model fit (colored lines), suggesting good agreement between the two. Notice also that overlap between the negative and positive component is small which bodes well for efforts to distinguish seronegative from seropositive samples. To further investigate the implications of the analyses, Figure S4 shows the estimated probability of infection as function of antibody concentration. Here, the probability of infection calculated as the estimated positive density (at a certain concentration) divided by the sum of the positive and negative densities (at that concentration) [1]. The figure shows that, in the absence of information on age-specific prevalence, the estimated probability of infection is close to 0 for concentrations of −1 (log(AU)/mL) and lower, and close to 1 at concentrations of 0 (log(AU)/mL) and higher.

**Figure S4.**
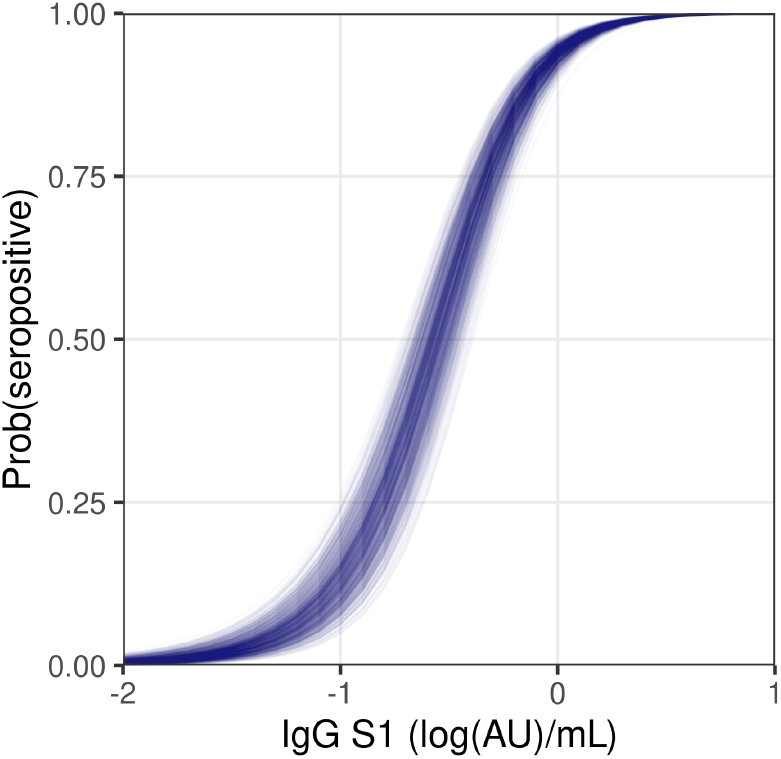
Estimated probability of seropositivity. Shown are estimated probabilities of seropositivity as function of the (log-transformed) antibody concentration. No weighting for prevalence is applied. Shown are 1, 000 samples from the posterior distribution.

In a next step we estimated the probability of seropositivity for each of the *n* = 6, 813 samples. Here we weighted the posterior seropositive density by the posterior prevalence, and the posterior seronegative density by 1 minus the posterior prevalence, and applied the same procedure as in Figure S4. The figure shows that for the majority of samples (6, 722), the posterior median for the probability of infection is either low (*<* 0.05, 6, 437 samples) or high (*>* 0.95, 285 samples), indicating that only for a small minority of samples (*<* 100) classification would not be straightforward. This is a robust result that also holds when using less informative priors or when including a random effect at the municipality level (not shown). It is due to the clear separation of the negative and positive components in the analyses (Figure S3).

**Figure S5.**
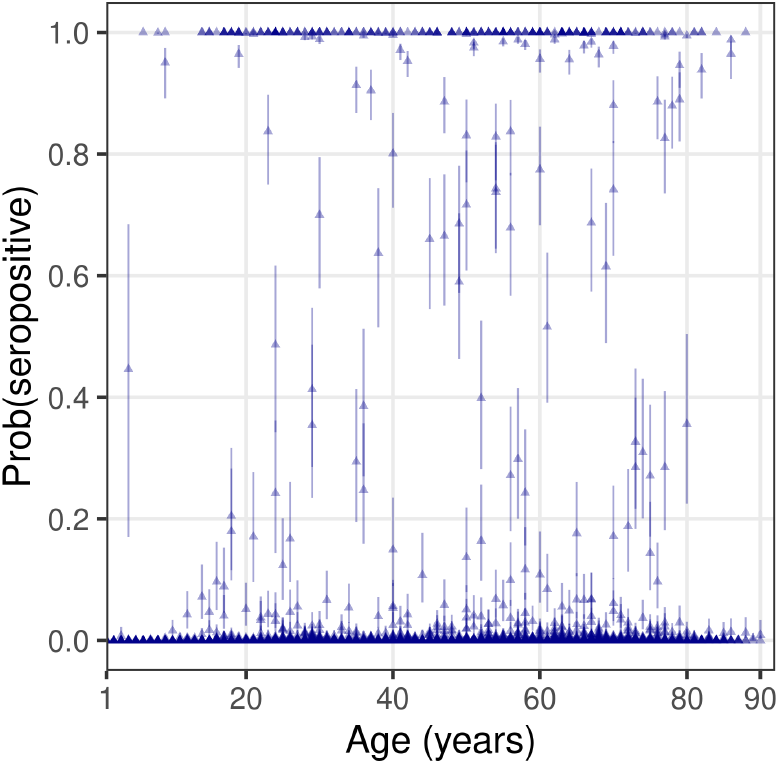
Estimated probability of seropositivity. Shown are estimated probabilities of seropositivity for each of the 6, 813 samples as function of age. Estimates are weighted with age-specific prevalence. Dots and whiskers represent posterior medians and 95% credible intervals, respectively. Notice that the posterior probability of seropositivity (i.e. posterior median) is either very low (< 0.05) or very high (> 0.95) for the majority of samples (> 98%).

### 7 Binary classification

The above results show that for the majority of samples there is limited uncertainty as to whether they should be classified as seronegative or seropositive. Therefore, we feel confident that reliable binary classification of the samples is feasible. Here, we investigate the optimal cut-off value for such binary classification, and associated test characteristics (sensitivity and specificity).

For a given cut-off, the proportion of the negative distribution with concentrations higher than the cut-off defines specificity of the test (high proportion implies low specificity), while the proportion of the positive distribution with concentrations lower than the cut-off defines sensitivity of the test. Technically, both sensitivity and specificity are calculated using cumulative density functions of the negative (specificity) and positive distributions (sensitivity) [1]. Figure S6 shows the test characteristics and the Youden index (*Se* + *Sp* − 1) as function of the cut-off. For low values of the cut-off, sensitivity of the test is high, at the price of a low specificity. Conversely, at high values of the cut-off, specificity of the test is high, at the price of low sensitivity. At intermediate values both sensitivity and specificity are reasonably high, and the Youden index is maximal.

**Figure S6.**
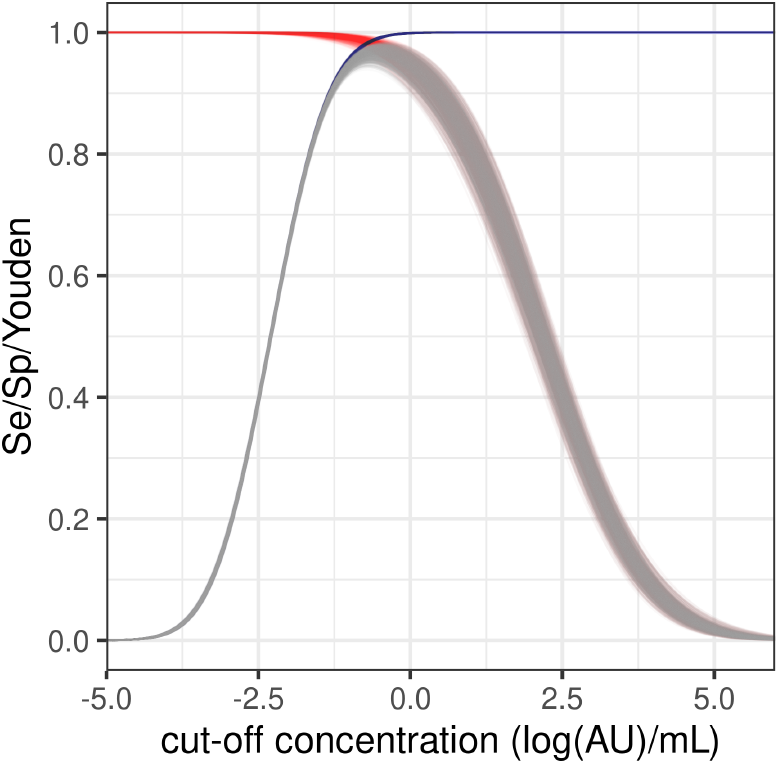
Sensitivity, specificity, and Youden index. Shown are the estimated sensitivity (red), specificity (blue), and Youden index (gray, superposed on top of sensitivity and specificity) as function of the cut-off concentration for seropositivity. Shown are 1, 000 samples from the posterior distribution.

**Table S3.** Test characteristics for cut-off that maximizes the Youden index or that selects for high test specificity (*Sp* = 0.999). Shown are posterior medians with 95% credible intervals.

| Scenario | cut-off (95%CrI) | $Se$ (95%CrI) | $Sp$ (95%CrI) | Youden (95%CrI) |
| --- | --- | --- | --- | --- |
| Youden | -0.56 (-0.67, -0.44) | 0.979 (0.965, 0.987) | 0.989 (0.985, 0.993) | 0.97 (0.95, 0.98) |
| Sp | 0.04 (0.0, 0.08) | 0.943 (0.910, 0.966) | 0.999 | 0.94 (0.91, 0.97) |

In Table S3 we show test characteristics for two specific scenarios. The first takes cut-offs that maximize the Youden index. Here, the estimated optimal cut-off is -0.56 (95%CrI: -0.67--0.44) and the estimated maximal Youden index is 0.97 (94%CrI: 0.95-0.98). This cut-off, however, is not useful in practice as expected seroprevalence is low (*<* 10%), and control of the false positive rate is more important than control of the false negative rate. Therefore, in a second scenario we aimed at a specificity of 0.999. Such specificity can be reached with the test, at a cut-off of 0.04 and a sensitivity of 0.943 (which is really good for such specificity!). In the following and in the main text we have opted for a cut-off of 0.04.

Figure S7 presents the results as a Receiver Operating Characteristic (ROC) diagram (blue lines), together with true and false positive rates at the cut-off of 0.04 (red dots). Variation in the false positive rate is minimal (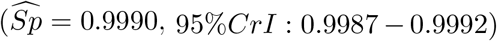, 95%*CrI*: 0.9987 − 0.9992), while estimated sensitivity is still high (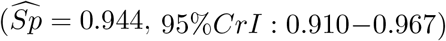, 95%*CrI*: 0.910−0.967). Estimated Youden index is 0.94 (95%*CrI*: 0.91−0.97).

**Figure S7.**
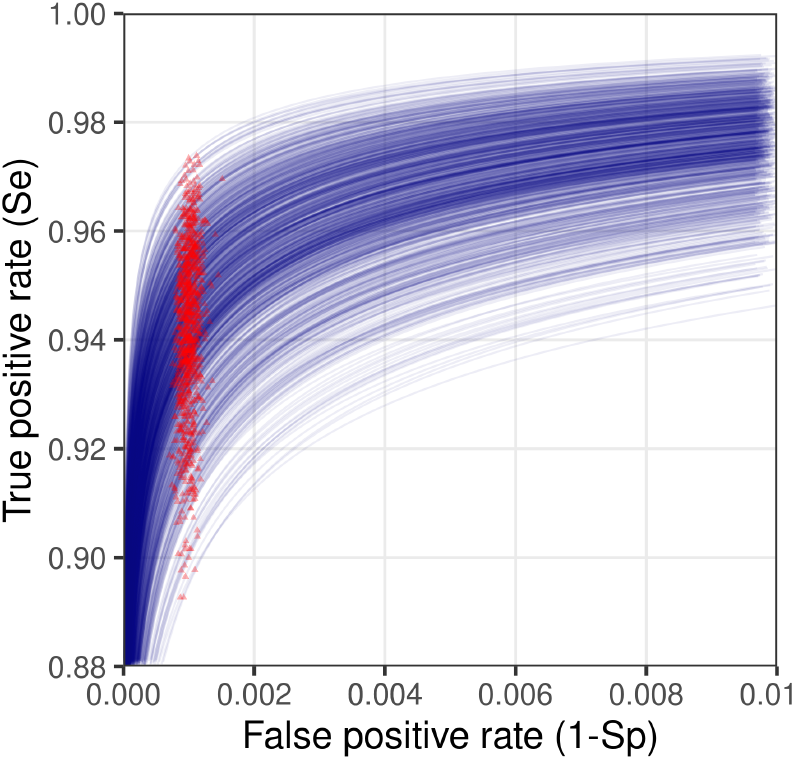
Receiver operator characteristic (ROC) diagram. Shown are the false positive rates (1 − Sp) and true positive rates (Se) for 1, 000 samples from the posterior distribution (blue). Also shown are the false and true positive rates for cut-off of 0.04 (log(AU)/mL) (1.04 AU/mL)(red).

Finally, Figure S8 shows the posterior distribution of test sensitivity at a cut-off of 0.04 (log(AU)/mL). Mean and standard deviation of the distribution are 0.942 and 0.0151, respectively. These values can be incorporated in Rogan-Gladen-type corrections for estimating true prevalence from observed apparent prevalence in binary classification [10, 11].

**Figure S8.**
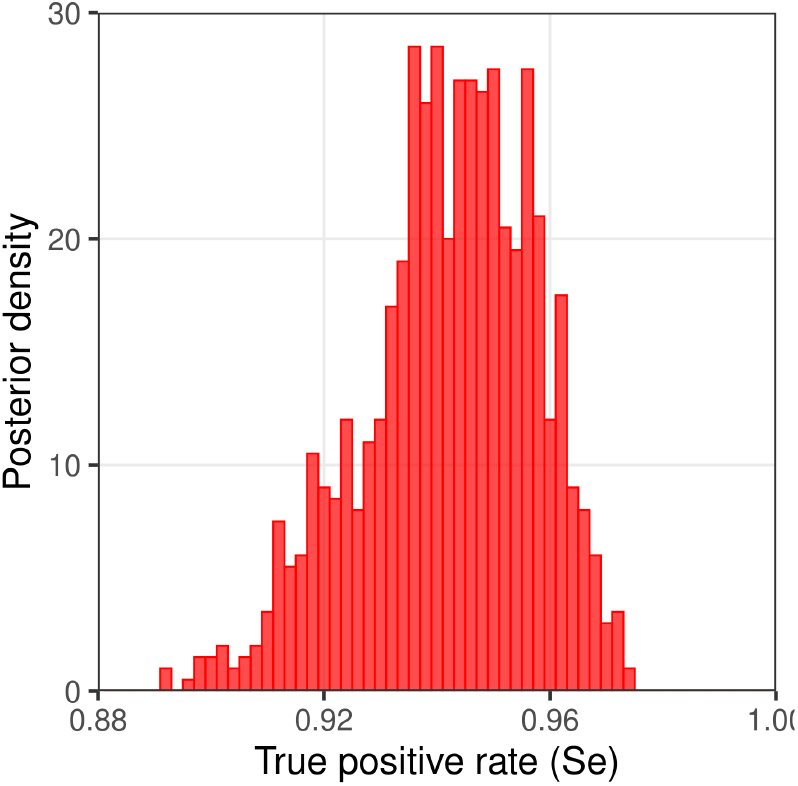
Posterior distribution of the true positive rate (sensitivity) when the cut-off is set at 0.04 (log(AU)/mL) (1.04 AU/mL). Shown is a histogram of 1, 000 samples from the posterior distribution. Mean and standard deviation of the distribution are 0.942 and 0.0151, respectively.

### 8 Logistic regression

The main text provides main results and interpretation of the analyses with logistic regression using the binary classification described in the above. Below we provide additional results on the regional estimates of seroprevalence (Figure S9), as well as the age-specific estimates of the unadjusted odd ratios for seropositivity derived from the univariable model (Figure S10).

**Figure S9.**
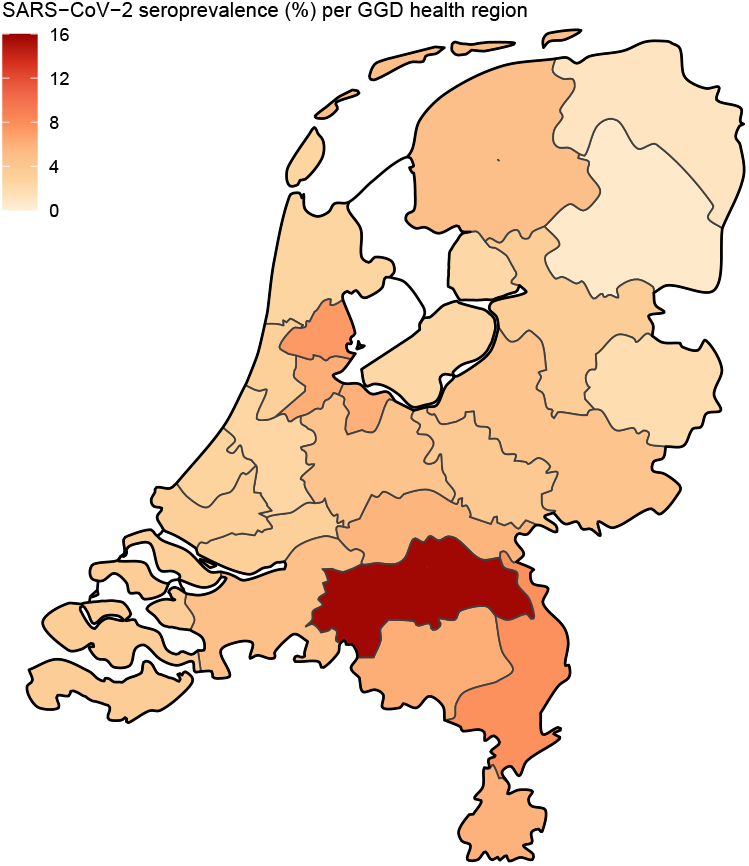
Estimates of seroprevalence by municipal health service area. Shown are estimates of (random effect) logistic regression based on the above binary classification, including sample weights and Rogan-Gladen bias correction [10].

**Figure S10.**
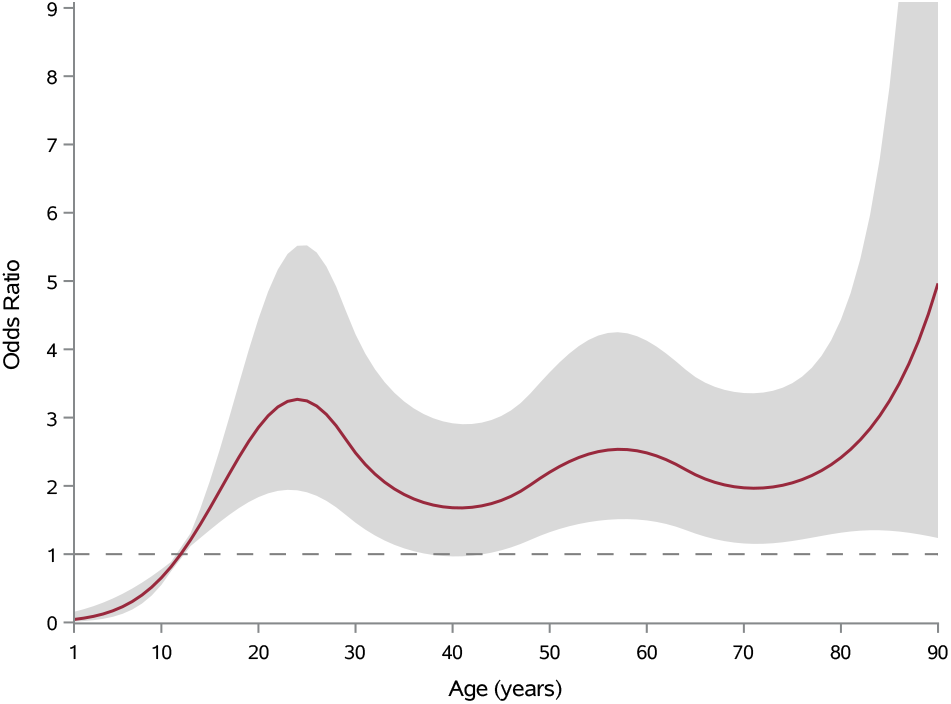
Estimates of the unadjusted odd ratios for seropositivity as function of age (see main text for adjusted odds ratios). The estimate is based on (random-effects) univariable logistic regression. Also shown is the 95% confidence envelope. Reference age is 12 years (odds ratio = 1).

